# Behavioral Changes During The Covid-19 Pandemic: Results Of A National Survey in Singapore

**DOI:** 10.1101/2020.08.06.20169870

**Authors:** Victoria JE Long, Jean CJ Liu

## Abstract

**Introduction:** As part of infection control measures for COVID-19, individuals have been encouraged to adopt both preventive (e.g., handwashing) and avoidant behavioural changes (e.g., avoiding crowds). In this study, we examined whether demographics predicted the likelihood that a person would adopt these behaviours in Singapore.

**Materials and Methods:** 1145 participants responded to an online survey conducted between 7 March - 21 April 2020. As part of the survey, we collected demographic information and asked participants to report which of 17 behaviour changes they had undertaken because of the outbreak. We ran regression models to predict, using demographic information: (1) the total number of behavioural changes undertaken, (2) the number of preventive changes undertaken, and (3) the number of avoidant changes undertaken. Finally, we sought to identify predictors of persons who: (4) declared that they had not undertaken any of these measures following the outbreak.

**Results:** Females and those who were younger adopted more preventive behaviours: whereas females were more likely to increase handwashing frequency, younger individuals were more likely to wear face masks prior to legislation. Females and those who were married adopted more avoidant behaviours, with both groups avoiding crowded areas and staying home more than usual. Females also voluntarily reduced physical contact, whereas those who were married chose outdoor venues and relied on online shopping.

**Conclusion:** Our characterisation of behavioural changes provides a baseline for public health advisories. Moving forward, local health authorities can focus their efforts to encourage segments of the population who do not readily adopt infection control measures against COVID-19.

---

Amidst the global outbreak of coronavirus disease 2019 (COVID-19), Singapore reported its first case on 23 January 2020^1^. Subsequently, the local ‘Disease Outbreak Response System Condition’ level was raised to Orange on 7 February 2020 when community transmission began. At this juncture, the government started to emphasise the role that individuals had to play by adopting health-protective behaviours^2^.

In an infectious disease outbreak such as the COVID-19 pandemic, individual-level health-protective behaviours can be classified into: (i) preventive behaviours - measures that can prevent transmission (e.g., hand-washing), and (ii) avoidant behaviours - measures that decrease contact with other individuals (e.g., avoiding crowded areas)^3^. As COVID-19 is believed to be transmitted primarily through contact or droplet transmission^4^, these measures can be effective in reducing the spread of the virus - particularly when pharmacological interventions are limited^5,6^.

For risk communication, it is useful to understand what characteristics predict whether an individual adopts health-protective behaviours. This allows public health messaging to be targeted, improving compliance in groups that may not do so as readily. For example, in the previous outbreak of severe acute respiratory syndrome (SARS), preventive and avoidant behaviours were more likely to be adopted by: women, older individuals, and those with higher education levels^3^. In the current COVID-19 outbreak, health-protective behaviours have been observed amongst individuals who perceive a higher risk of infection, higher disease severity, or who are afraid of getting infected^7–9^. However, demographic predictors have differed between populations studied: whereas age and gender were linked to behavioural changes in South Korea, these associations were not found in the United Kingdom^8,9^. Further, no demographic predictors were identified in a study in the United States, while gender - but not age - predicted behavioural changes in a cross-country survey^10,11^. This heterogeneity suggests that the uptake of health-protective behaviours may be context-specific during the COVID-19 pandemic, owing - perhaps - to heterogeneity in the risks of infection or risk of severe illness between countries.

In light if this context-specificity of demographic studies, we conducted a large-scale survey to examine how demographics predict the uptake of health-protective behaviours in Singapore to characterise our local population. Our study was conducted across March-April 2020, a period when the country saw a rapid increase in COVID-19 cases (from 138 cases at the start of the study, to 9125 cases at the end of the survey period).

## Methods

### Study design and population

From 7 March-21 April 2020, 1145 participants responded to an online survey on COVID-19. As the inclusion criteria, participants: (1) were aged ≥21 years old, and (2) had lived in Singapore for ≥2 years. Given public health concerns, participants were recruited online - via advertisements placed in community chat-groups (e.g., Facebook and WhatsApp groups for residential estates, universities, and workplaces) or via paid Facebook advertisements targeting Singapore-based users. The study was approved by the Yale-NUS College Ethics Review Committee (#2020-CERC-001), and participants gave written consent in accordance with the Declaration of Helsinki. The questions reported in this study were part of a larger 20-minute survey exploring: behavioural and psychological responses to COVID-19, sources from which participants received COVID-19 news, and psychological wellbeing (https://osf.io/pv3bj)^12^.

### Predictor variables

As predictors, participants reported the following demographic details: gender, ethnicity, religion, country of birth, marital status, education, house type, and household size. As behavioural changes may be influenced by the local COVID-19 situation, we also recorded the total number of local cases reported to date, and whether the country was locked down when the survey was done (computed based on the survey time-stamp).

### Outcome variables

As the key outcome variables, participants indicated which of 17 health-protective behaviours they had voluntarily undertaken because of the pandemic (by indicating ‘yes’ or ‘no’ for each behaviour). Based on prior research^3^, we investigated 3 preventive behaviours, asking participants whether they had: (1) washed their hands more frequently, (2) used hand sanitisers and/or (3) wore a mask in public (prior to legislation). Additionally, we investigated 14 possible avoidant behaviours, whether participants had: (1) avoided crowded areas, (2) reduced physical contact, (3) stayed home more than usual, (4) distanced from people with flu symptoms, (5) voluntarily changed travel plans, (6) missed or postponed social events, (7) avoided visiting hospitals and/or healthcare settings, (8) chose outdoor over indoor venues, (9) distanced from people with recent travel to outbreak countries, (10) distanced from people with possible contact with COVID-19 cases, (11) avoided places where COVID-19 cases were reported, (12) stored up more household and/or food supplies, (13) relied more on online shopping (prior to shop closures), and/or (14) avoided public transport. Across the 17 items, we assigned a score of ‘1’ for ‘yes’ responses, and these were summed to create three scores: the total number of behavioural changes adopted (out of 17), the total number of preventive behaviours adopted (subscale score out of 3), and the total number of avoidant behaviours adopted (subscale score out of 14).

Finally, we included as a separate item the following statement: “I did not take any additional measures” (yes/no response). This question allowed us to identify participants who had not made any behavioural changes as a function of COVID-19 - a group that may be of higher risk for transmission.

### Statistical analyses

To describe participants’ demographic characteristics, survey responses were summarized with counts. As the primary analysis, we then ran a linear regression model with the total number of behavioural changes as the outcome measure, and participant demographics as predictors (age, gender, ethnicity, religion, country of birth, marital status, education, house type, household size, the total number of local cases reported to date, and whether the country was locked down at the time of survey completion) [Model 1]. Given that prior research distinguished preventive and avoidant behaviours^3,13^, we repeated the linear regression model with the total number of preventive behaviours [Model 2], and the total number of avoidant behaviours as outcomes [Model 3]. Finally, we ran a logistic regression model to identify - using the same demographic predictors - individuals who had made no behavioural changes [Model 4].

For linearity, the number of local COVID-19 cases was log-transformed prior to regression analyses. For each regression model, the type 1 family-wise error rate was controlled at 0.05 through Bonferroni correction (Bonferroni-adjusted alpha level of 0.05/23 predictors=0.002). All statistical analyses were conducted using R (Version 4.0) and STATA (Version 12.0).

## Results

### Response rate

Of 1390 individuals who clicked the survey link, 1145 (82.4%) provided informed consent and participated in the survey. A further 192 (16.77%) participants were excluded from statistical analyses as they did not complete the primary outcome measures (on behavioural changes).

As shown in Table 1, the final sample of 953 participants was comparable to the resident Singapore population in: the proportion of Singapore citizens, marital status, and household size (≤10% difference). However, the pool of respondents had a greater representation of females (65.1% vs. 51.1%), university graduates (72.7% vs. 32.4%), and persons of no religion (28.0% vs. 18.5%) or Christian belief (36.2% vs. 18.8%). Conversely, there was a reduced representation of participants who lived in 1-3 room public housing flats (6.7% vs. 23.7%). Survey respondents were also more likely to be of Chinese ethnicity than persons in the general population (87.0% vs. 74.3%) (14).

**Table 1.**
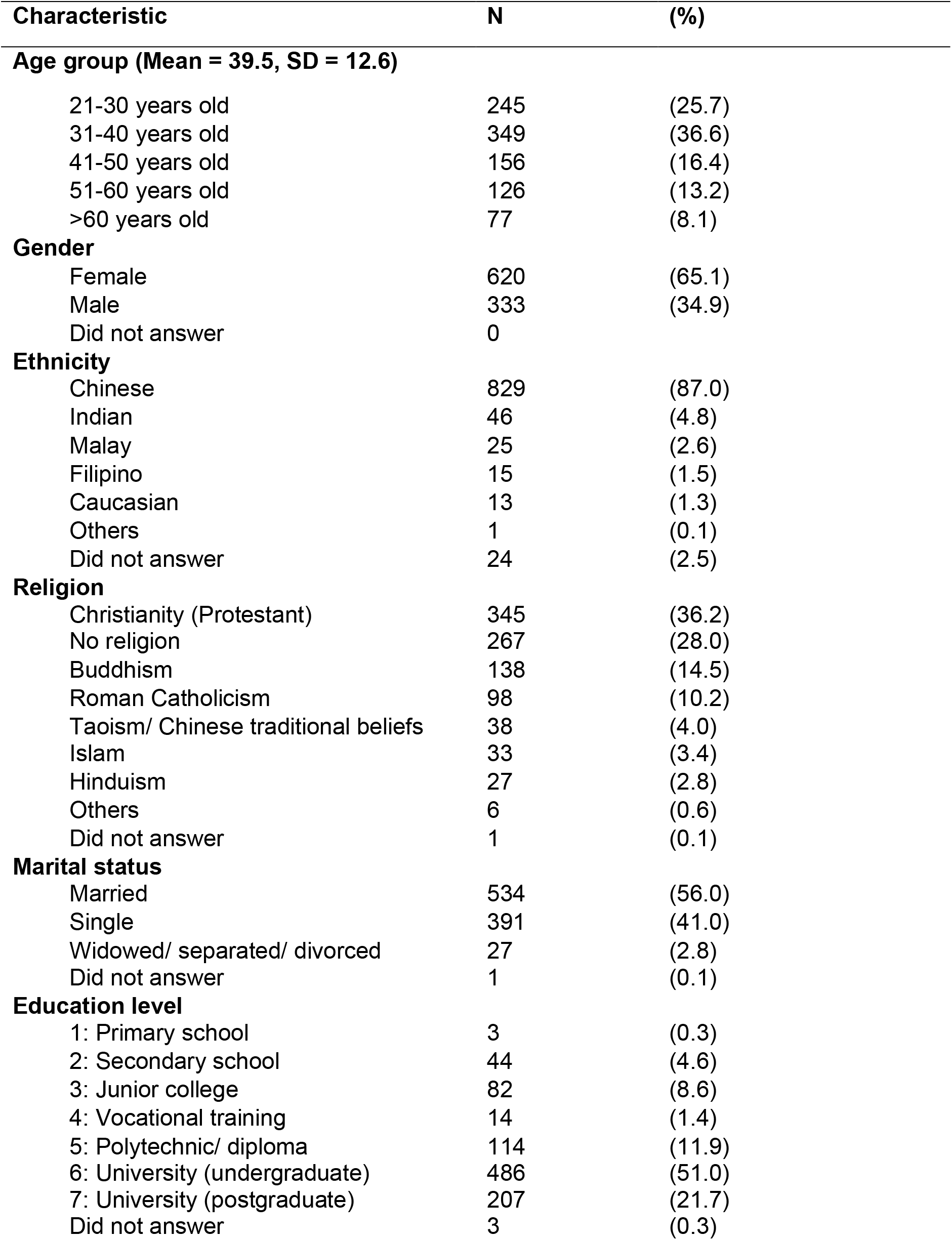
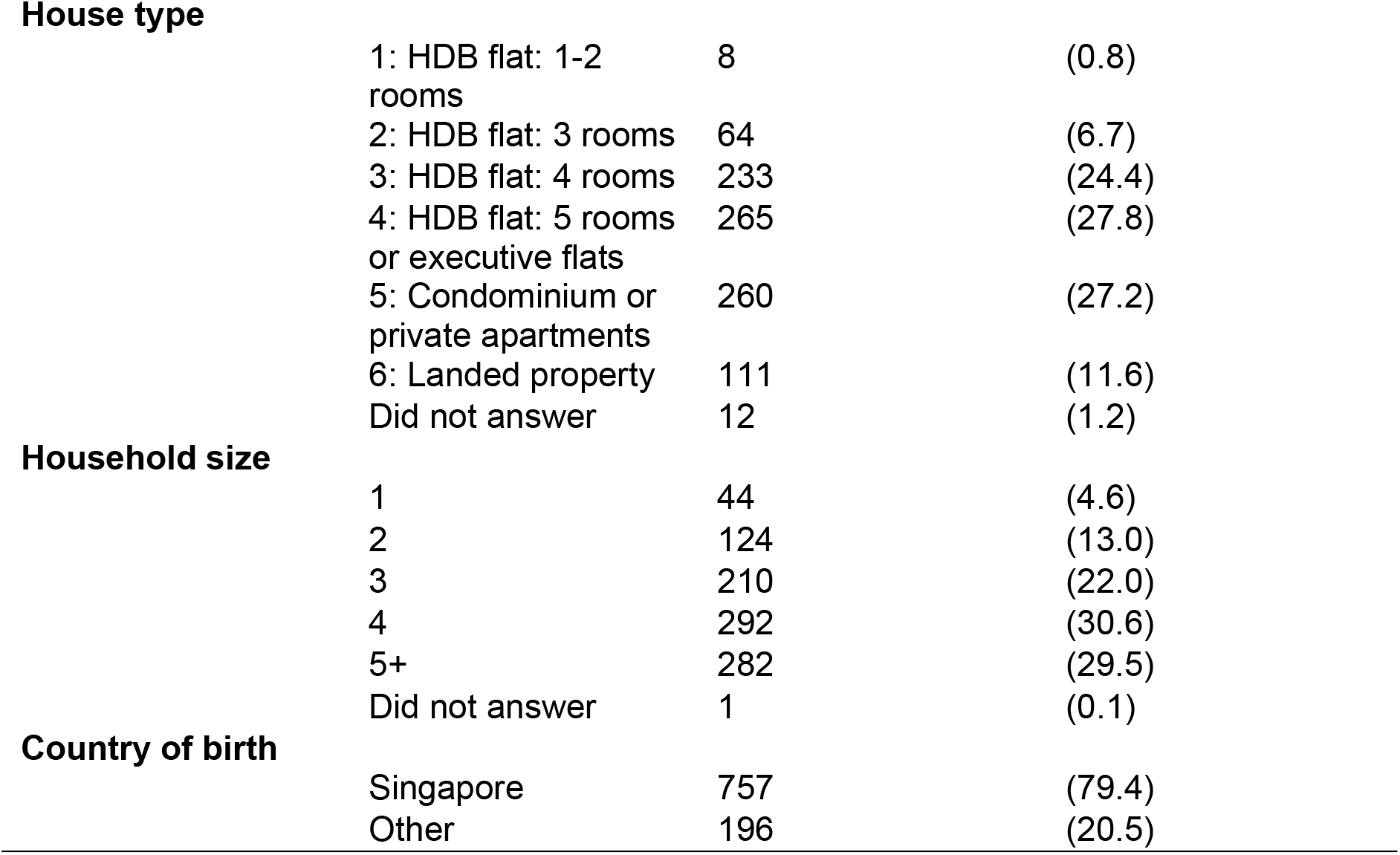
Baseline demographics of participants.

| Characteristic | N | (%) |
| --- | --- | --- |
| **Age group (Mean = 39.5, SD = 12.6)** |  |  |
| 21-30 years old | 245 | (25.7) |
| 31-40 years old | 349 | (36.6) |
| 41-50 years old | 156 | (16.4) |
| 51-60 years old | 126 | (13.2) |
| >60 years old | 77 | (8.1) |
| **Gender** |  |  |
| Female | 620 | (65.1) |
| Male | 333 | (34.9) |
| Did not answer | 0 |  |
| **Ethnicity** |  |  |
| Chinese | 829 | (87.0) |
| Indian | 46 | (4.8) |
| Malay | 25 | (2.6) |
| Filipino | 15 | (1.5) |
| Caucasian | 13 | (1.3) |
| Others | 1 | (0.1) |
| Did not answer | 24 | (2.5) |
| **Religion** |  |  |
| Christianity (Protestant) | 345 | (36.2) |
| No religion | 267 | (28.0) |
| Buddhism | 138 | (14.5) |
| Roman Catholicism | 98 | (10.2) |
| Taoism/ Chinese traditional beliefs | 38 | (4.0) |
| Islam | 33 | (3.4) |
| Hinduism | 27 | (2.8) |
| Others | 6 | (0.6) |
| Did not answer | 1 | (0.1) |
| **Marital status** |  |  |
| Married | 534 | (56.0) |
| Single | 391 | (41.0) |
| Widowed/ separated/ divorced | 27 | (2.8) |
| Did not answer | 1 | (0.1) |
| **Education level** |  |  |
| 1: Primary school | 3 | (0.3) |
| 2: Secondary school | 44 | (4.6) |
| 3: Junior college | 82 | (8.6) |
| 4: Vocational training | 14 | (1.4) |
| 5: Polytechnic/ diploma | 114 | (11.9) |
| 6: University (undergraduate) | 486 | (51.0) |
| 7: University (postgraduate) | 207 | (21.7) |
| Did not answer | 3 | (0.3) |

|  |  |  |  |
| --- | --- | --- | --- |
| **House type** |  |  |  |
|  | 1: HDB flat: 1-2 rooms | 8 | (0.8) |
|  | 2: HDB flat: 3 rooms | 64 | (6.7) |
|  | 3: HDB flat: 4 rooms | 233 | (24.4) |
|  | 4: HDB flat: 5 rooms or executive flats | 265 | (27.8) |
|  | 5: Condominium or private apartments | 260 | (27.2) |
|  | 6: Landed property | 111 | (11.6) |
|  | Did not answer | 12 | (1.2) |
| **Household size** |  |  |  |
|  | 1 | 44 | (4.6) |
|  | 2 | 124 | (13.0) |
|  | 3 | 210 | (22.0) |
|  | 4 | 292 | (30.6) |
|  | 5+ | 282 | (29.5) |
|  | Did not answer | 1 | (0.1) |
| **Country of birth** |  |  |  |
|  | Singapore | 757 | (79.4) |
|  | Other | 196 | (20.5) |

### An overview of COVID-19 behaviour changes

On the whole, participants adopted a mean of 8.01 (SD=3.78) behavioural changes owing to the COVID-19 pandemic. This corresponded to a mean of 2.14 (SD=0.81) preventive measures, and 5.87 (SD=3.44) avoidant measures. Only 29 participants (3.04%) reported that they had not changed their behaviours at all.

### Predicting behavioural change: regression models

In our first regression model, we sought to predict the total number of behaviour changes based on participant demographics (Table 2). We first observed that behavioural changes tracked the local COVID-19 situation: namely, as the number of local cases increased, individuals adapted their behaviours in response (b=3.03, t(913)=3.96, p<0.001). Having controlled for local transmission, gender emerged as a significant predictor, with females adopting an average of 0.14 more changes than males (t(913)=-4.49, p<0.001). Being married was also associated with a higher number of health-protective behaviours than being single (b=1.09, t(913)=3.52, p<0.001).

In our second and third models, we examined whether demographic predictors differed for preventive vs. avoidant behaviours. In terms of demographics, while the adoption of preventive behaviours was predicted by gender (b=-0.241, t(913)=-4.33, p<0.001) and age (b=-0.008, t(913)=-3.11, p=0.001), the adoption of avoidant behaviours was predicted by gender (b=-0.902, t(913)=-3.90, p<0.001) and marital status (being married vs. being single; b=0.973, t(913)=3.45, p<0.001).

Finally, in our fourth model, we found that no demographic predictor significantly identified the small proportion of individuals who had not undertaken any measures on account of COVID-19 (all *p* > Bonferroni-adjusted alpha of 0.002).

**Table 2.** Predicting behavioural changes during the COVID-19 outbreak.

|  | Outcome<sup>a</sup> |  |  |  |
| --- | --- | --- | --- | --- |
|  | (1)<br>Total number of<br>behaviours | (2)<br>Preventive<br>behaviours | (3)<br>Avoidant<br>behaviours | (4)<br>No behaviour<br>changes adopted |
| **Age** | -0.002 (0.011) | -0.008 (0.002)* | 0.005 (0.010) | 0.041 (0.018) |
| **Gender**<br>(base = female) |  |  |  |  |
| Male | -1.14 (0.254)* | -0.241 (0.055)* | -0.902 (0.231)* | 0.757 (0.411) |
| **Ethnicity**<br>(base = Chinese) |  |  |  |  |
| Indian | -1.20 (1.39) | -0.743 (0.304) | -0.463 (1.26) | -31.4 (268) |
| Malay | -1.01 (0.949) | -0.397 (0.208) | -0.620 (0.863) | -13.4 (172) |
| Filipino | 1.04 (1.10) | 0.430 (0.242) | 0.617 (1.00) | -29.4 (327) |
| Caucasian | -0.004 (1.05) | -0.318 (0.796) | 0.313 (0.962) | -15.6 (292) |
| Others | 1.42 (3.63) | -2.21 (0.085) | 3.64 (3.30) | -14.6 (1080) |
| **Religion**<br>(base = no religion) |  |  |  |  |
| Christianity<br>(Protestant) | 0.795 (0.390) | 0.172 (0.085) | 0.623 (0.354) | -0.542 (0.518) |
| Buddhism | 0.291 (0.632) | 0.037 (0.138) | 0.254 (0.574) | -1.20 (0.816) |
| Roman<br>Catholicism | 0.800 (1.34) | 0.360 (0.294) | 0.440 (1.21) | -13.7 (1.10) |
| Taoism/<br>Chinese<br>traditional<br>beliefs | 1.63 (1.18) | -0.034 (0.259) | 1.32 (1.07) | -0.555 (1.10) |
| Islam | 0.647 (0.463) | -0.034 (0.101) | 0.681 (0.421) | 15.5 (172) |
| Hinduism | 0.209 (0.310) | -0.014 (0.400) | 0.223 (0.282) | 31.3 (268) |
| Others | -4.07 (1.82) | -0.629 (0.400) | -3.44 (1.65) | 2.08 (1.26) |
| **Marital status**<br>(base = single) |  |  |  |  |
| Married | 1.09 (0.309)* | 0.117 (0.067) | 0.973 (0.281)* | -0.739 (0.552) |
| Widowed/<br>separated/<br>divorced | -0.495 (0.433) | 0.009 (0.094) | -0.504 (0.393) | -0.750 (0.850) |
| **Education level** | 0.003 (0.093) | -0.005 (0.020) | 0.009 (0.084) | -0.342 (0.126) |
| **House type** | 0.141 (0.113) | 0.014 (0.024) | 0.126 (0.102) | -0.064 (0.185) |
| **Household size** | -0.013 (0.114) | 0.047 (0.025) | -0.060 (0.103) | 0.124 (0.208) |
| **Country of birth**<br>(base = Singapore) |  |  |  |  |
| Other | -0.730 (0.338) | -0.152 (0.074) | -0.578 (0.307) | -0.169 (0.589) |
| **Lockdown**<br>(base = yes) |  |  |  |  |
| No lockdown | -1.76 (0.017) | -0.621 (0.161)* | -1.138 (0.670) | -0.043 (1.21) |
| **Number of local**<br>**COVID-19 cases**<br>**(log transformed)** | 3.03 (0.612)* | 0.531 (0.134)* | 2.50 (0.556)* | -0.151 (0.986) |
| **$R^2$** | 0.123 | 0.091 | 0.124 | 0.144 |

<sup>a</sup>Data reported as beta estimates (standard error)
\*Indicates significance at $p < 0.002$ (following Bonferroni corrections).

### Follow-up exploratory analyses: Understanding which behavioural changes differed as a function of gender, marital status, and age

To better understand the pattern of results, we conducted post-hoc chi-square tests to identify which behavioural changes differed as a function of gender and marital status (for all behaviours), and as a function of age (for preventive behaviours). As these were exploratory analyses, the type 1 decision-wise error rate was controlled at 0.05 (uncorrected).

#### Gender

As shown in Figure 1, females were more likely than males to: 1) wash their hands more frequently, χ^2^(1, N=953)=22.17, p<0.001; 2) avoid crowded areas, χ^2^(1, N=953) = 11.83, p=0.001; 3) reduce physical contact, χ^2^(1, N=953)=9.28, p=0.002, and 4) stay home more than usual, χ^2^(1, N=953)=9.79, p=0.002.

#### Marital status

As shown in Figure 2, marital status was significantly associated with: avoiding crowded areas, χ^2^(2, N=952)=26.29, p<0.001; 2) staying home more than usual, χ^2^(2, N=952)=28.09, p<0.001; 3) choosing outdoor over indoor areas, χ^2^(2, N=952)=33.04, p<0.001; and 4) relying more on online shopping, χ^2^(2, N=952)=26.37, p<0.001. In each case, single participants were least likely to adopt these behaviours than those who were not single (married, widowed, separated, or divorced).

#### Age

Finally, wearing a mask in public differed between age groups, χ^2^(4,N=953)=33.32, p<0.001), with participants aged 21-30 most likely to adopt this behaviour (Figure 3).

#### Sensitivity analyses

As the chi-square analyses examined behavioural changes as a function of one predictor at a time (either gender, marital status, or age), we repeated our analyses by regressing each behavioural change against the full set of demographics (per Models 1-4 previously). Our conclusions did not change, and the full table of chi-square analyses and regression results are reported in the Appendix.

**Figure 1.**
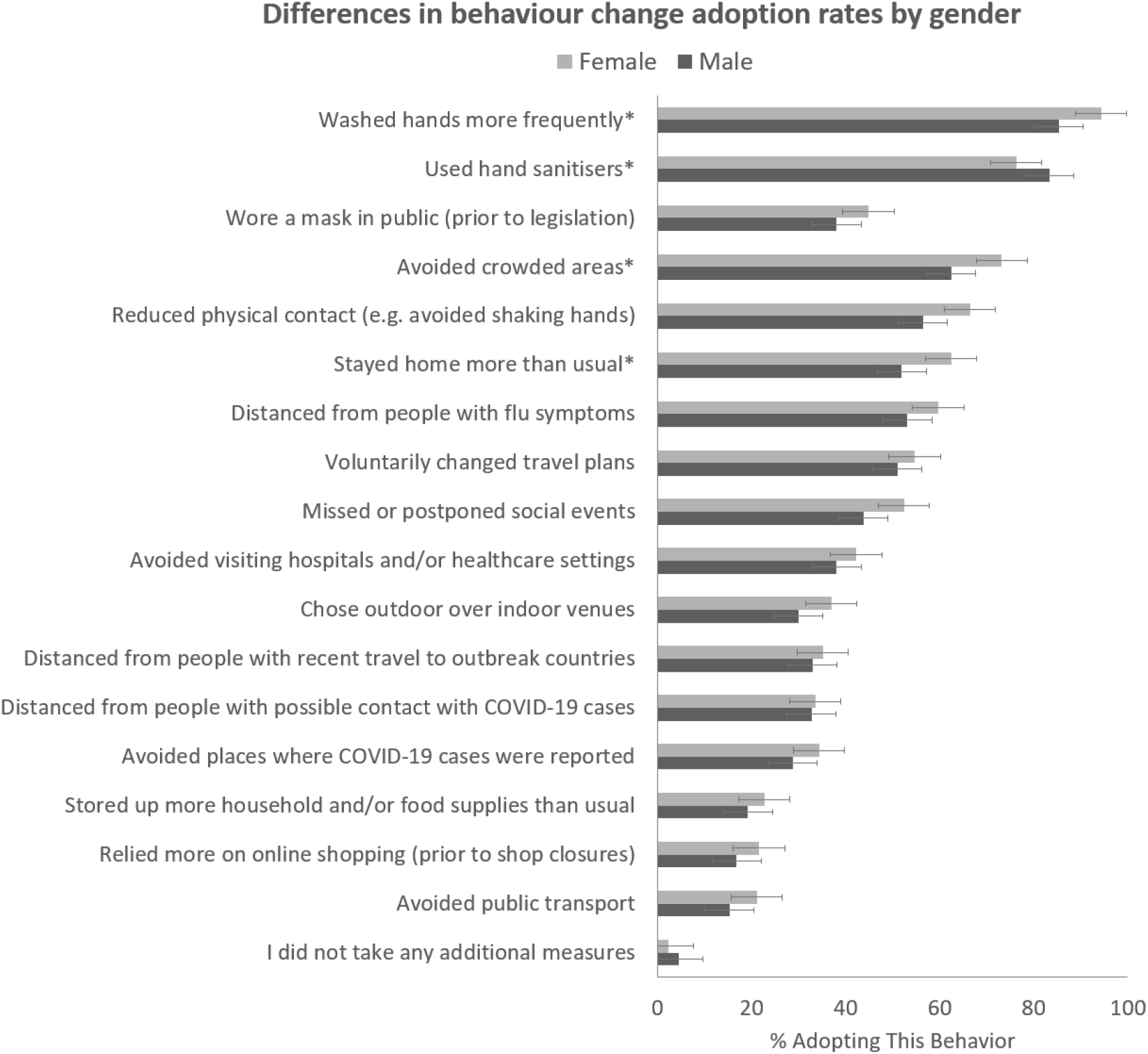
Uptake of COVID-19 infection control measures as a function of gender. Asterisks indicate significance at *p* < 0.002 (following Bonferroni corrections), and horizontal lines represent the 95% confidence intervals.

**Figure 2.**
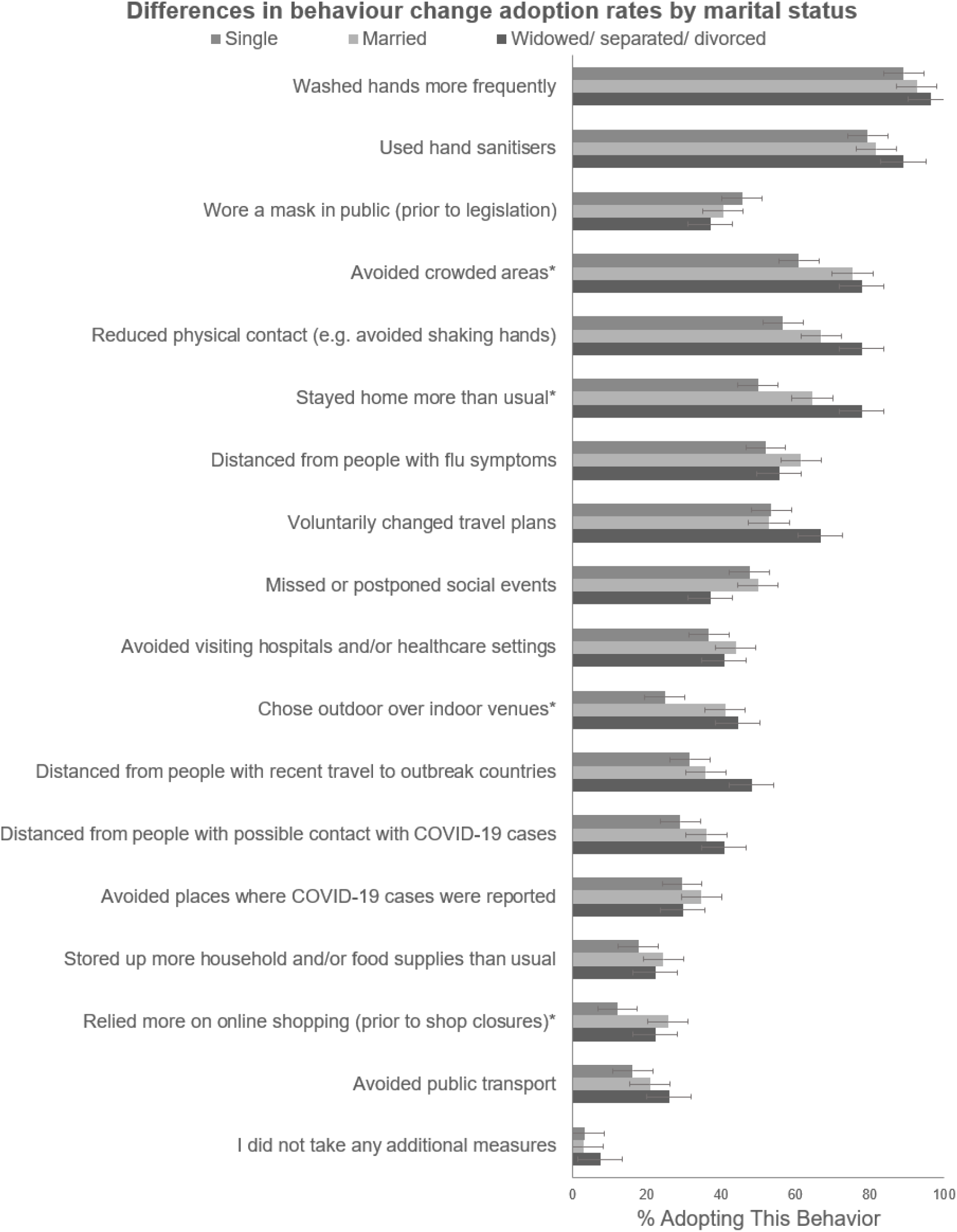
Uptake of COVID-19 infection control measures as a function of marital status. Asterisks indicate significance at *p* < 0.002 (following Bonferroni corrections), and horizontal lines represent the 95% confidence intervals.

**Figure 3.**
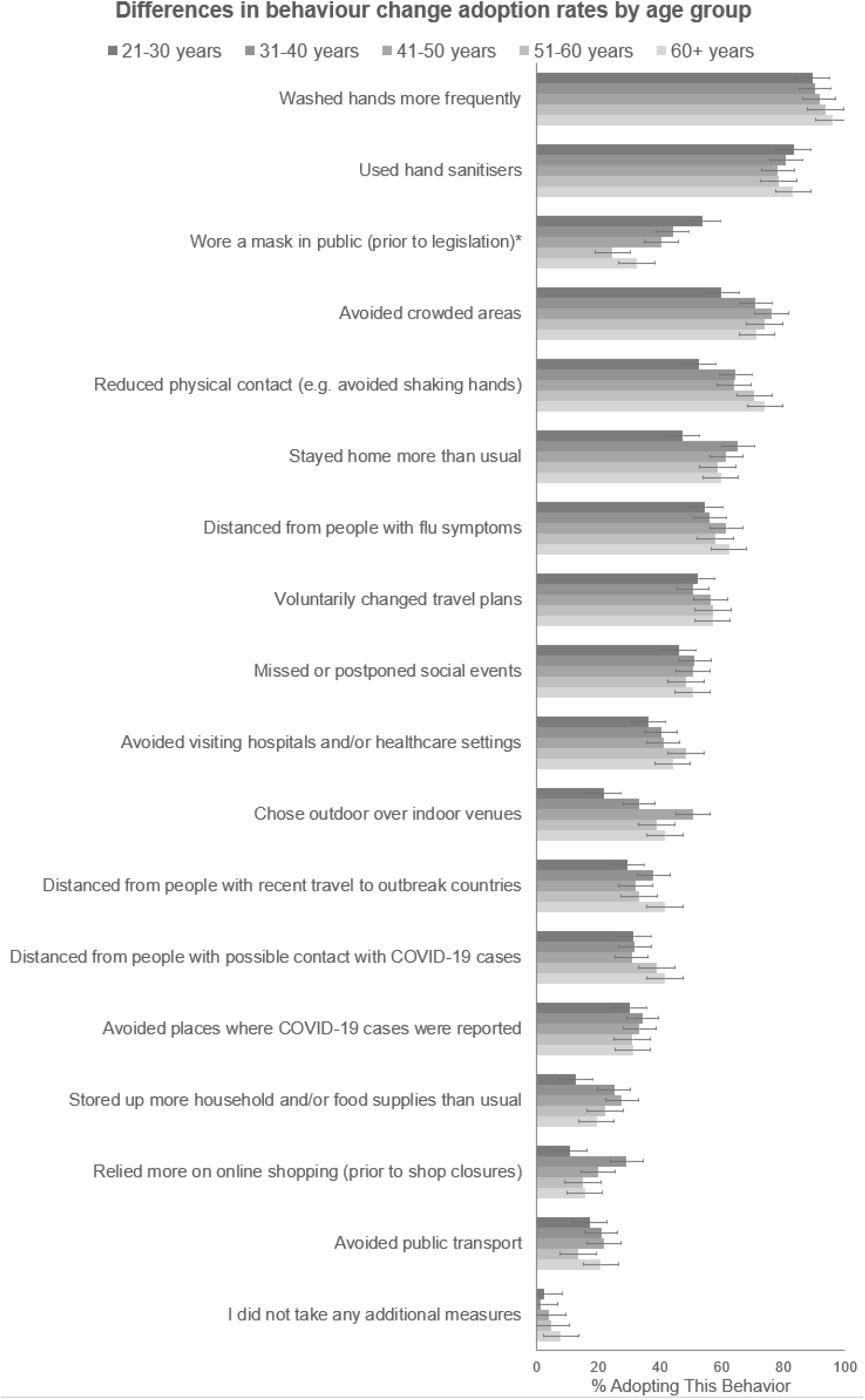
Uptake of COVID-19 infection control measures as a function of age group. Asterisks indicate significance at *p* < 0.002 (following Bonferroni corrections), and horizontal lines represent the 95% confidence intervals.

## Discussion

In this study, we documented for the first time how residents in Singapore have adapted their behaviours to minimize COVID-19 transmission. The large majority of participants (97%) have undertaken at least one infection control measure, with participants reporting an average of 8 lifestyle changes owing to the pandemic. As might be expected, behavioural changes increased with the number of COVID-19 cases reported locally.

In terms of demographic predictors, health-preventive measures were most likely to be adopted by females and those who were married. When we distinguished between preventive (e.g. hand washing) and avoidant (e.g. avoiding crowded areas) behaviours, age emerged as an additional predictor for avoidant behaviours, with youths most likely to adopt mask-wearing.

Collectively, our results on gender and marital status replicate findings from previous infectious disease outbreaks^3,15^ and the current COVID-19 pandemic (based on both an international and a South Korean sample^8,10^). These findings echo a broader pattern of risk that has emerged in epidemiological research, whereby being female and being married has been linked to the reduced risk of disease and of all-cause mortality^16^. Adding to this body of research, our findings highlight how being willing to adopt health-promoting behaviours during a pandemic may contribute to the resilience of these demographic groups.

Departing from prior research and popular belief, however, we found that age was inversely related to the take-up of preventive behaviours. In particular, younger adults in our survey were more likely to wear masks than older adults, even before legislation stipulating that masks had to be worn in public. This finding is remarkable for several reasons. First, during SARS, older adults had been more likely to perform a range of preventive behaviours including mask-wearing, handwashing, respiratory hygiene, the using of utensils, and washing after touching contaminated surfaces (3). Second, during the current outbreak, several high-profiled events (e.g., coronavirus parties hosted by students) have resulted in the belief that youths are least likely to care about the outbreak, and thereby most likely to ignore infection control measures^17,18^. Indeed, the Director-General of the World Health Organization released a statement telling youths that they were “not invincible”, that “the virus could put (them) in hospital for weeks, or even kill (them)”^19–21^.

Rather than finding young persons to take on risky behaviours, however, we observed instead that this demographic group was most associated with mask-wearing. While this finding is counter-intuitive, it is in line with recent Hong Kong research whereby elderly participants - rather than the young - were least likely to worry about getting infected, and thus least likely to adopt protective behaviours^22^. Additionally, young persons’ ready adoption of mask-wearing may reflect a general willingness to embrace change and innovation, since mask-wearing had not previously been a norm in Singapore (as it had in countries like Japan^23^).

### Policy implications

Moving forward, our findings may contribute to the public health strategy in several ways. First, throughout the pandemic, government agencies have repeatedly noted how individuals have ignored official advisories^24,25^. This phenomenon has been so widespread that the individuals have been nick-named ‘covidiots’ in the popular press - a portmanteau of coronavirus and idiot^26,27^. Beyond ‘naming and shaming’, however, our research highlights characteristics that may predict noncompliance. This, in turn, will allow risk communication to be targeted.

On the other hand, our findings also highlight which demographic groups may be most likely to respond when the government launches a new infection control measure (for example, SafeEntry or TraceTogether for contact tracing). Extrapolating from our research, these initiatives - if perceived to be health-protective - may be adopted first by females and those who are married. Correspondingly, the two demographic groups may be ideal for pilot trials or as advocates for the behaviours.

### Limitations

In making these recommendations, we note that our study has several limitations. First, we relied on participants’ self-reports, which may be vulnerable to recollection biases. Future research will need to explore whether our findings translate to actual behavioural changes during the pandemic. Second, although our survey methodology captured behavioural changes at one particular time-point, the recommendation of infection control measures is a moving target. In the case of mask-wearing, for example, official advisories changed from masks not being needed, to being encouraged, to finally being mandated (as of 14 April 2020)^28^. Correspondingly, further research is needed to examine whether our findings continue to hold even as official advisories change.

### Conclusions

In conclusion, we conducted the first Singapore-based study of behavioural changes during the COVID-19 pandemic. Although the scale of this crisis has been unprecedented and many uncertainties remain, many of our findings reinforce longstanding patterns of how demographic characteristics can pre-dispose an individual to disease - in this case, via the uptake of measures that can minimize COVID-19 infection. Moving forward, our findings provide a template by which official messaging can be tailored for health promotion.

## Data Availability

Data will be made available at the request to the corresponding author.

## Acknowledgements

This research was funded by a grant awarded to JCJL from the JY Pillay Global Asia Programme (grant number: IG20-SG002). The authors gratefully acknowledge Saw Young Ern and Edina Tan for assisting with manuscript preparation.

## Appendix

**Appendix 1:** Proportion of respondents who adopted each behaviour change, and chi-square results for each behaviour change by gender, marital status and age group

|  | Total %<br>(SE) | Chi-square |  |  |
| --- | --- | --- | --- | --- |
|  |  | Gender | Marital status | Age group<sup>1</sup> |
| **Preventive** |  |  |  |  |
| Washed hands more frequently | 91.18<br>(0.91) | 22.17* | 4.75 | 4.67 |
| Used hand sanitisers | 80.90<br>(1.27) | 7.08 | 2.17 | 2.30 |
| Wore a mask in public (prior to legislation) | 42.49<br>(1.60) | 3.98 | 3.08 | 33.32* |
| **Avoidant** |  |  |  |  |
| Avoided crowded areas | 69.46<br>(1.49) | 11.83* | 26.29* | - |
| Reduced physical contact (e.g. avoided shaking hands) | 62.95<br>(1.56) | 9.28* | 13.39 | - |
| Stayed home more than usual | 58.76<br>(1.59) | 9.79* | 28.09* | - |
| Distanced from people with flu symptoms | 57.39<br>(1.60) | 3.77 | 8.85 | - |
| Voluntarily changed travel plans | 53.41<br>(1.61) | 1.14 | 2.00 | - |
| Missed or postponed social events | 49.42<br>(1.62) | 6.37 | 2.65 | - |
| Avoided visiting hospitals and/or healthcare settings | 40.81<br>(1.59) | 1.52 | 6.28 | - |
| Chose outdoor over indoor venues | 34.52<br>(1.54) | 4.57 | 33.04* | - |
| Distanced from people with recent travel to outbreak countries | 34.41<br>(1.53) | 0.06 | 7.29 | - |
| Distanced from people with possible contact with COVID-19 cases | 33.26<br>(1.52) | 0.43 | 6.10 | - |
| Avoided places where COVID-19 cases were reported | 32.42<br>(1.51) | 3.01 | 3.68 | - |
| Stored up more household and/or food supplies than usual | 21.51<br>(1.33) | 1.59 | 6.87 | - |
| Relied more on online shopping (prior to shop closures) | 19.93<br>(1.29) | 3.12 | 26.37* | - |
| Avoided public transport | 19.09<br>(1.27) | 4.73 | 4.49 | - |

\*Indicates significance at $p < 0.002$ (following Bonferroni corrections).
<sup>1</sup>Age groups were separated as: 21-30 years old, 31-40 years old, 41-50 years old, 51-60 years old and above 60 years old.

**Appendix 2:**
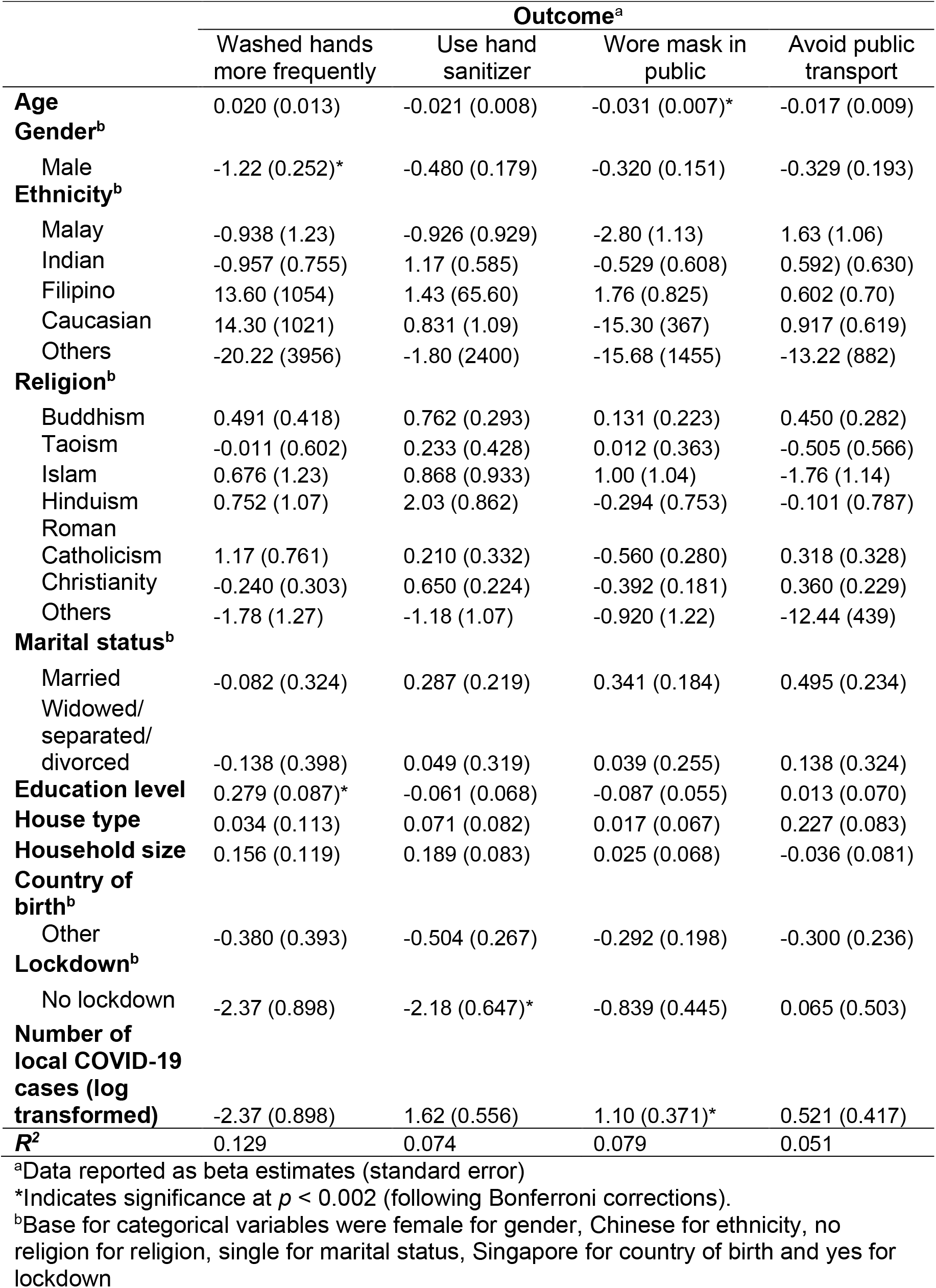
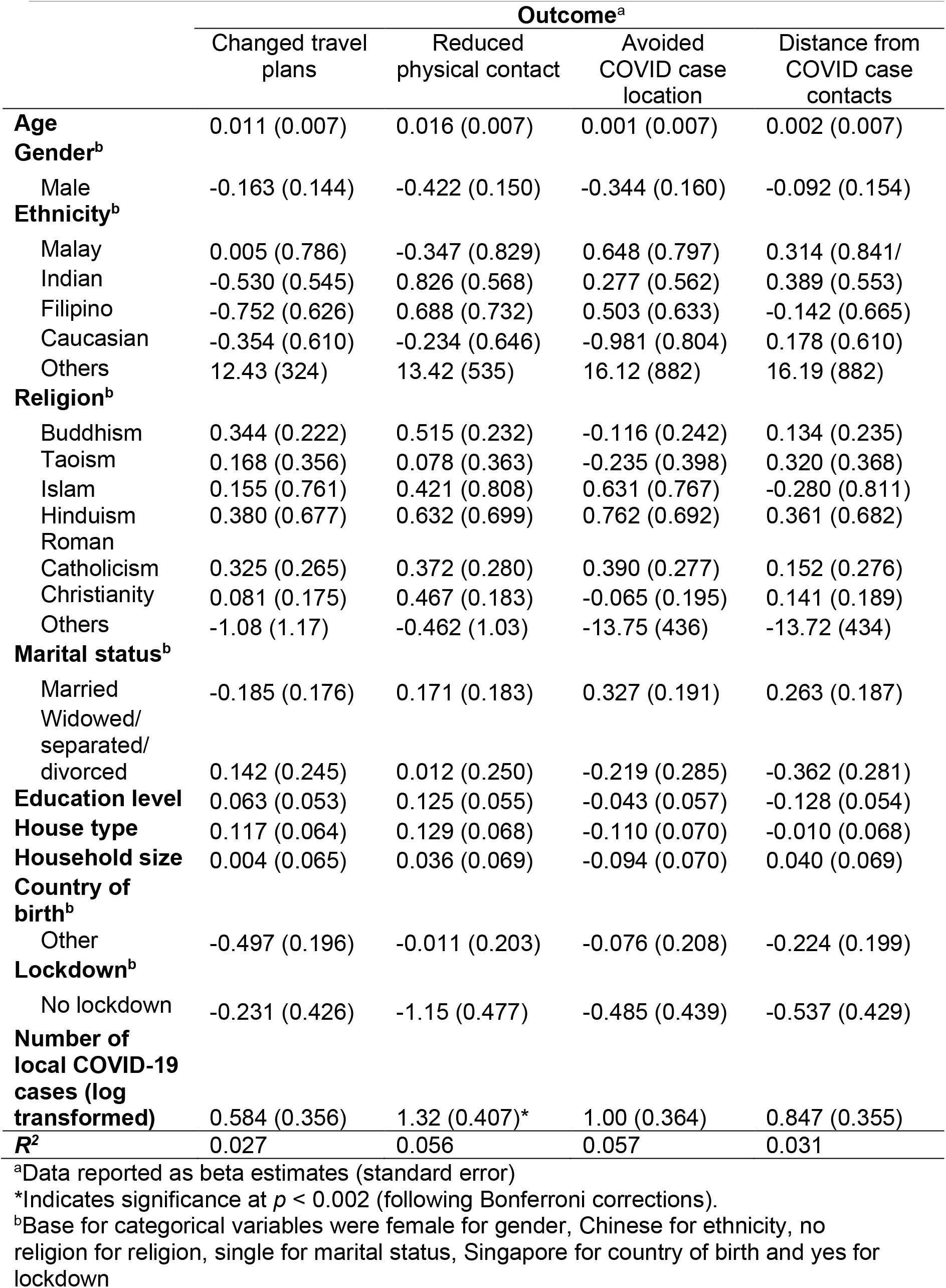
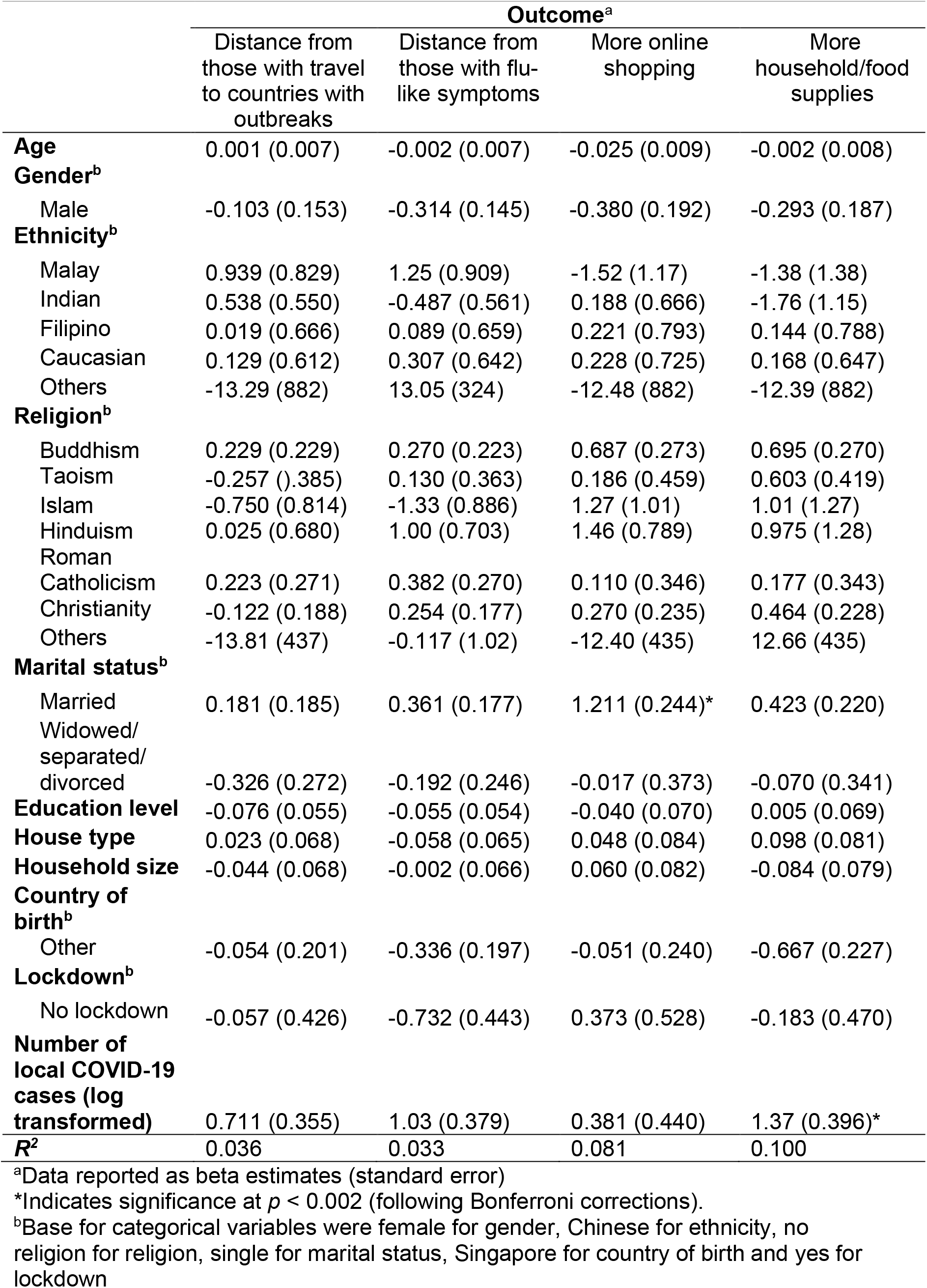
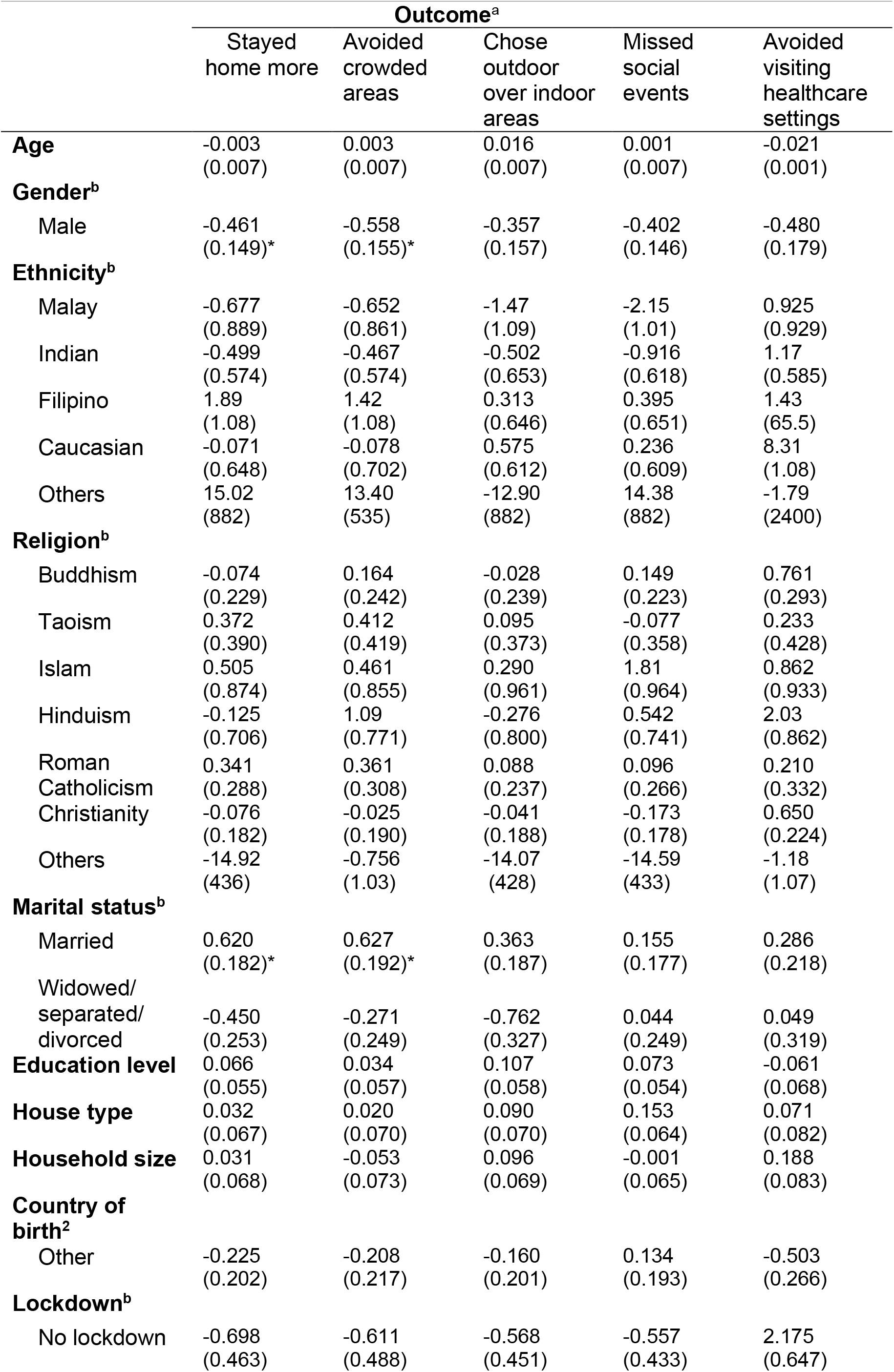
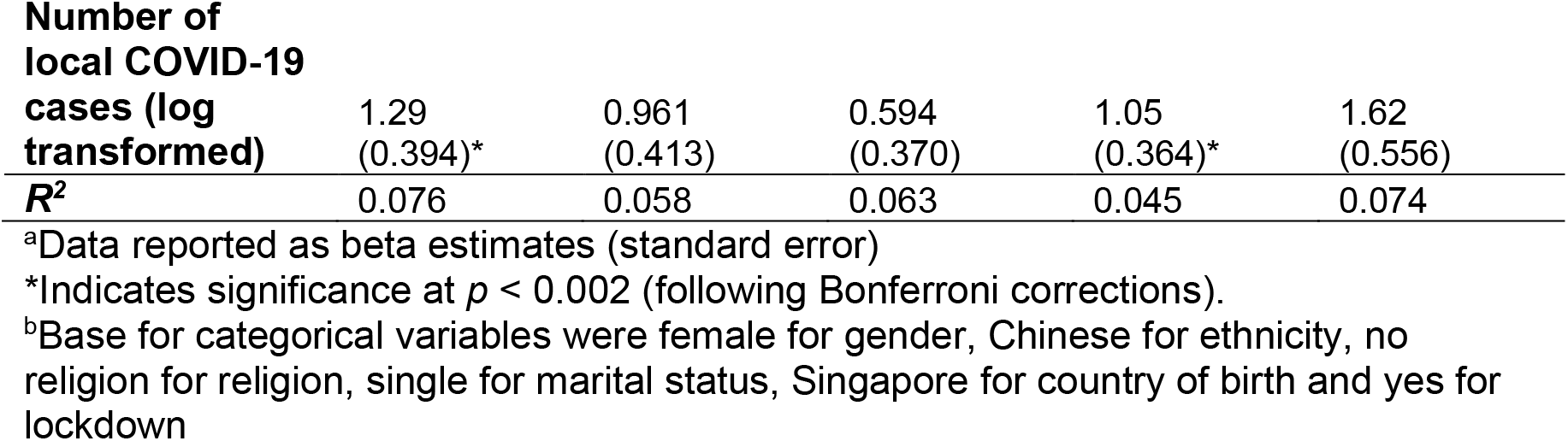
Predicting each of the 17 behaviour changes

## Notes

### Competing Interest Statement

The authors have declared no competing interest.

### Funding Statement

This research was funded by a grant awarded to the corresponding author from the JY Pillay Global Asia Programme (grant number: IG20-SG002).

### Author Declarations

The study was approved by the Yale-NUS College Ethics Review Committee (#2020-CERC-001), and participants gave written consent in accordance with the Declaration of Helsinki.

